# Trajectories of depressive symptoms among vulnerable groups in the UK during the COVID-19 pandemic

**DOI:** 10.1101/2020.06.09.20126300

**Authors:** Philipp Frank, Eleonora Iob, Andrew Steptoe, Daisy Fancourt

## Abstract

**Objective:** The coronavirus disease 2019 (COVID-19) pandemic has affected many aspects of the human condition, including mental health and psychological wellbeing. This study examined trajectories of depressive symptoms (DST) over time among vulnerable individuals in the UK during the COVID-19 pandemic.

**Methods:** The sample consisted of 51,417 adults recruited from the COVID-19 Social Study. Depressive symptoms were measured on seven occasions (21st March - 2nd April), using the Patient Health Questionnaire (PHQ-9). Sociodemographic vulnerabilities included non-white ethnic background, low socio-economic position (SEP), and type of work (keyworker versus no keyworker). Health-related and psychosocial vulnerabilities included pre-existing physical and mental health conditions, experience of psychological and/or physical abuse, and low social support. Group-based DST were derived using latent growth mixture modelling and multivariate logistic regression models were fitted to examine the association between these vulnerabilities and DSTs. Model estimates were adjusted for age, sex, and suspected COVID-19 diagnosis.

**Results:** Three DSTs were identified: low [N=30,850 (60%)] moderate [N=14,911 (29%)], and severe [N=5,656 (11%)] depressive symptoms. DSTs were relatively stable across the first 6 weeks of lockdown. After adjusting for covariates, experiences of physical/psychological abuse (OR 13.16, 95% CI 12.95-13.37), pre-existing mental health conditions (OR 13.00 95% CI 12.87-13.109), pre-existing physical health conditions (OR 3.41, 95% CI 3.29-3.54), low social support (OR 12.72, 95% CI 12.57-12.86), and low SEP (OR 5.22, 95% CI 5.08-5.36) were significantly associated with the severe DST. No significant association was found for ethnicity (OR 1.07, 95% 0.85-1.28). Participants with key worker roles were less likely to experience severe depressive symptoms (OR 0.66, 95% 0.53-0.80). Similar but smaller patterns of associations were found for the moderate DST.

**Conclusions:** People with psychosocial and health-related risk factors, as well as those with low SEP seem to be most vulnerable to experiencing moderate or severe depressive symptoms during the COVID-19 pandemic.

## INTRODUCTION

Due to the rapid spread of the coronavirus disease 2019 (COVID-19), large numbers of people across the UK have been urged to stay at home (“lockdown”) for potentially significant periods of time. It is already evident that the COVID-19 pandemic and its related containment measures have profound implications for many aspects of society ^1^. Up to now, much focus has been placed on investigating the incidence and mortality rates of COVID-19, the pathogenesis of the virus, its adverse effects on physical health, and its growing impact on the global economy. But there is growing awareness of the implications of the pandemic for mental health ^2^. The consequences of the COVID-19 pandemic are likely to occur against the backcloth of elevated prevalence of mental health problems in the UK. The widespread experience of COVID-19-related stressors (e.g. loss of employment, illness or death of a relative), reduced access to mental health services, and frequent concerns about mental and physical health in the general population highlight the importance of identifying who is most at risk and how their experiences are evolving as the pandemic continues.

Previous studies on former epidemics and pandemics (e.g. the severe acute respiratory syndrome epidemic (SARS) in 2003 or the 2014 Ebola outbreak) suggest a plausible increase in mental health problems across the wider population, including symptoms of post-traumatic stress, confusion, anger and depression ^2,3^. However, it is unclear how mental health problems are manifesting in more vulnerable groups during the current pandemic, such as in people with pre-existing physical and/or mental health conditions, people with experiences of physical and psychological abuse, and people of lower socio-economic position (SEP). Prior to the COVID-19 outbreak, the prevalence of depression was estimated at 4-5% in the general population ^4,5^, with considerably higher rates observed in individuals with distinct health-related, sociodemographic and psychosocial vulnerabilities ^6^. For example, the prevalence of depression in people with chronic obstructive pulmonary disease (COPD) has been placed at 27% ^7^, type II diabetes 18-20% ^8,9^, myocardial infarction 20% ^10^, cancer 13-17% ^11,12^, and stroke patients at 29-33% ^13,14^. Furthermore, meta-analytic evidence indicates significantly elevated rates of depression in women who experienced intimate partner violence (27%) ^15^, and almost a two-fold increased risk of depression in people with lower SEP ^16^.

Evidence already suggests that pre-existing health inequalities may be strongly reflected in the current COVID-19 pandemic. For example, recent mortality statistics indicate that the severe acute respiratory syndrome coronavirus 2 (SARS-CoV-2), the virus that causes COVID-19, may be particularly detrimental to members of Black, Asian and minority ethnic (BAME) groups, with approximately 19% of UK COVID-19-related hospital deaths occurring in members of the BAME community ^17^. In addition, key workers (e.g. frontline healthcare and social care staff) may be particularly vulnerable to developing symptoms of emotional distress. According to a study of the psychosocial effects associated with SARS in a sample of 510 hospital workers, 29% of health-care staff reported elevated symptoms of emotional distress ^18^. The COVID-19 pandemic and related containment measures are also likely to accentuate social isolation and feelings of loneliness ^1^. Notably, these factors are themselves strongly associated with the incidence, severity and progression of negative mental health outcomes, including depression, anxiety-related disorders, and suicide ^19^. Other COVID-19 related stressors such as loss of employment, financial hardship, overcrowded households, and diminished access to social support networks may further contribute to disease burden, nationally and globally, by increasing levels of distress and reducing social opportunities relevant to both mental and physical wellbeing ^20^.

Given the possibility that specific sociodemographic, psychosocial, and health-related characteristics may make some people particularly vulnerable to poorer mental health, an immediate research priority is to investigate the psychological wellbeing in these groups, and to understand how it evolves over time as the pandemic continues. Therefore, the aim of the present study was to explore trajectories of depressive symptoms (DST) among vulnerable individuals in the UK during the COVID-19 pandemic.

## METHOD

### Study design and participants

The COVID-19 Social Study is an ongoing large panel study of adults aged 18 years and older residing in the UK. The study was established on 21^st^ March 2020, using online weekly data collection to explore the psychological and social experiences of adults during the COVID-19 pandemic. The sample was recruited using three primary approaches. First, snowballing was used, including promoting the study through existing networks and mailing lists (including large databases of adults who had previously consented to be involved in health research across the UK), print and digital media coverage, and social media. Second, more targeted recruitment was undertaken focusing on (i) individuals from a low-income background, (ii) individuals with no or few educational qualifications, and (iii) individuals who were unemployed. Third, the study was promoted via partnerships with third sector organisations to vulnerable groups, including adults with pre-existing mental health conditions, older adults, carers, and people experiencing domestic violence or abuse. The study was approved by the UCL Research Ethics Committee [12467/005] and all participants gave informed consent.

The present analysis focused on participants recruited between 21^st^ March and 4^th^ May 2020. All duplicate email addresses were removed prior to the sample size being calculated and only individuals who provided fully completed interviews on at least one wave were included. This resulted in a final analytical sample of 51,417 participants. Although participant selection was based on a multi-stage non-random sampling approach, the sample was well-stratified across sociodemographic characteristics and all data were weighted to the proportions of gender, age, ethnicity, education and country of living obtained from the Office for National Statistics (ONS, 2018).

## Measures

### Outcome: Depressive symptoms

Depressive symptoms were measured using the Patient Health Questionnaire (PHQ-9), a validated screening tool for diagnosing depression in primary care ^21^. The questionnaire involves nine items asking about the frequency of experiencing common symptoms of major depressive disorder during the past week ^22^. Each item was answered on a 4-point Likert scale, ranging from “not at all” to “nearly every day”. Higher overall scores indicate more depressive symptoms, with scores of 0-4 suggesting minimal depression, 5-9 mild depression, 10-14 moderate depression, 15-19 moderately severe depression, and scores of 20-27 denoting severe depression ^23^.

### Exposures

#### a) Health-related vulnerabilities

##### (I) Pre-existing physical health conditions

Pre-existing physical health conditions were measured by asking participants whether they had been clinically diagnosed with (i) diabetes, (ii) high blood pressure, (iii) heart disease, (iv) lung disease (e.g. asthma or COPD), (v) cancer, and (vi) another clinically diagnosed chronic physical health condition. Each item was answered on a binary response scale (yes = 1, no = 0). For the purposes of the present analyses, a binary score was computed, distinguishing between the presence (at least one pre-existing physical health condition) and absence (no pre-existing physical health condition) of a clinically diagnosed physical illness.

##### (II) Pre-existing mental health conditions

Pre-existing mental health conditions were assessed by asking participants whether they had a clinical diagnosis of (i) depression, (2) anxiety, and/or (3) other mental health condition. Items were ranked on a binary response scale (yes = 1, no = 0). A summary score was computed, categorising responses into the presence (at least one clinically diagnosed health condition) versus absence (no pre-existing mental health conditions) of a pre-existing mental health condition.

#### b) Sociodemographic vulnerabilities

##### (I) Ethnicity

Participants were asked to provide information about their ethnic backgrounds. Ethnicity was categorised into two groups: White (i.e. British, Irish or other) versus non-White (i.e. Asian/Asian British, Black/Black British, White and Black/Black British, Mixed race, Chinese/Chinese British, Middle Eastern/Middle Eastern British, other ethnic group).

##### (II) Socio-economic position index

First, a continuous latent socio-economic position (SEP) index was computed using five indicators of SEP: (i) household income, (ii) employment status, (iii) education, (iv) household tenure, and (v) household overcrowding. Income was measured by self-reported annual household income. Responses were categorised as “< £16,000 a year”, “£16,000-£29,999 a year”, “£30,000-£59,999 a year”, “£60,000-89,999 a year”, and “£90,000 or more per year”. Employment was measured by asking participants to provide information about their employment status. Responses were grouped as “employed”, “inactive”, and “unemployed”. Education was assessed in reference to participants’ highest educational qualification (postgraduate, undergraduate, A-level or vocational, and GCSE or lower). Household tenure was categorised as “owned outright” versus “owned with mortgage / shared ownership” versus “rented”. Overcrowded households were defined as households with < 1 room per occupant. Next, we computed a binary SEP index variable by first dividing the continuous latent SEP variable into quartiles, and then categorising the highest quartile as “low SEP”.

##### (III) Key worker roles

Participants were asked whether they had currently fulfilled any of the government’s identified key worker roles (yes = 1, no = 0). Key workers included people with jobs that were deemed essential during the pandemic and included those in health and social care, education and childcare, key public services, local and national government, public safety and national security, transport, as well as utility workers. People were only classed as keyworkers if their role involved them leaving the home to carry out this work during the lockdown.

#### c) Psychosocial vulnerabilities

##### (I) Experience of psychological and/or physical abuse

Participants indicated whether during the last week they had been “physically harmed or hurt by someone else” or “bullied, controlled, intimidated, or psychologically hurt by someone else”. Responses were rated on a 4-point Likert scale ranging from “not at all” to “nearly every day”. A binary variable was created focusing on any response on either item that indicated any experience of abuse on at least one occasion (yes = 1, no = 0).

##### (II) Social support

Social support was measured using an adapted version of the six-item short form of Perceived Social Support Questionnaire (F-SozU K-6) ^24^. This includes six questions asking participants whether, during the past week, they had “experienced a lot of understanding and support from others”, “a very close person whose help they can always count on”, “people with whom they can spend time and do things together”, “friends and family who will take care of them if they get sick”, “people they can talk to without hesitation if they feel down”, and whether they could “easily borrow something from neighbours or friends if needed”. Each item is rated on a 5-point scale from “not true at all” to “very true”, with higher scores indicating higher levels of perceived social support. Minor adaptations were made to the language in the scale to make it relevant to experiences during COVID-19 (see sTable 1 for a comparison of changes, Supplement). A sum score was computed by adding up the mean scores (across all available waves) of each social support question per individual and dividing it into quartiles. We subsequently derived a binary social support variable, defining “low social support” as scores in the lowest quartile.

### Covariates

Covariates included age, gender, and the presence/absence of a (suspected) COVID-19 diagnosis.

### Statistical analysis

Group-based trajectories of depressive symptoms were estimated using latent growth mixture (LGM) modelling (Newsom, 2015). This data-driven modelling technique enables the identification of individuals with similar DSTs by computing classes (or trajectories) of average values within homogenous subgroups over time. Next, multivariate logistic regression models were fitted to examine the associations of sociodemographic, psychosocial, and health-related vulnerabilities with group-based DST. We first tested the individual effects of the vulnerabilities by fitting separate regression model for each exposure (Model 1). Second, we fitted a model including all exposures simultaneously to assess their mutually adjusted effects (Model 2). All estimates were adjusted for age, gender, and the presence/absence of a (suspected) COVID-19 diagnosis. The LGM models were fitted using Robust Maximum Likelihood Estimation to account for missing data and for the non-normal distribution of the depression scores. The results of all multivariate logistic regression analyses are presented as adjusted odds ratios (OR) with corresponding 95% confidence intervals (CI). Data management and descriptive analyses were conducted using R version 3.4.4. LGM and multivariate logistic regression analyses were performed using Mplus version 7. Further details about the LGM analysis and model fit can be found in the Supplement (sMethods, sTable 2).

## RESULTS

### Descriptive statistics

The characteristics of the study participants at the first assessment are reported in Table 1 (see sTable 3 and sTable 4 for the weighted and unweighted descriptive statistics at each wave, Supplement). The weighted sample was 51% female, and 12% of participants had a non-white ethnic background. There was a higher proportion of participants in the oldest age groups (32%) compared with the youngest groups (18%). 60% of participants were employed and 22% were undertaking key worker roles. In contrast, around 40% of the sample was inactive or unemployed. The majority of participants had GCSE or A-level qualifications, and around 30% also had degree qualifications. There was a higher proportion of participants in the low- and medium-income groups compared with the highest ones. A total of 8% of participants lived in overcrowded households. Almost 40% of participants had a pre-existing physical illness, 20% reported having at least one mental health problem, and almost 1 in 3 participants had moderate or severe symptoms of depression at the first assessment [see sFigure 1 (Supplement) for the average levels of depressive symptoms throughout the study]. 11% had experienced psychological or physical abuse on at least one occasion since lockdown began.

**Table 1.** Characteristics of the study participants (N=51,417)

| **Table 1. Characteristics of the study participants (N=51,417)** |  |
| --- | --- |
| **Sex** |  |
| Female | 26276 (51.1%) |
| Male | 25140 (48.9%) |
| **Age** |  |
| 18-29 | 9228 (17.9%) |
| 30-44 | 11972 (23.3%) |
| 45-59 | 13723 (26.7%) |
| 60+ | 16494 (32.1%) |
| **Ethnicity** |  |
| Non-white | 6145 (12.0%) |
| White | 45272 (88.0%) |
| **Employment status** |  |
| Employed | 30888 (60.1%) |
| Inactive | 19414 (37.8%) |
| Unemployed | 1115 (2.2%) |
| **Education** |  |
| Postgraduate | 6964 (13.5%) |
| Undergraduate | 10453 (20.3%) |
| A-Level or Vocational | 17362 (33.8%) |
| GCSE or Lower | 16638 (32.4%) |
| **Income** |  |
| N-Miss | 5308 |
| <£16k | 9704 (21.0%) |
| £16k - £30k | 12846 (27.9%) |
| £30k - £60k | 14434 (31.3%) |
| £60k - £90k | 5488 (11.9%) |
| >£90k | 3638 (7.9%) |
| **Overcrowding** |  |
| Yes | 4196 (8.2%) |
| **Tenure** |  |
| N-Miss | 179 |
| Own Mortgage | 15991 (31.2%) |
| Own Outright | 16016 (31.3%) |
| Rent | 19231 (37.5%) |
| **Socioeconomic position index <sup>a</sup>** |  |
| 1<sup>st</sup> quartile (=highest) | 8353 (16.2%) |
| 2<sup>nd</sup> quartile | 9525 (18.5%) |
| 3<sup>rd</sup> quartile | 16395 (31.9%) |
| 4<sup>th</sup> quartile (=lowest) | 17143 (33.3%) |
| **Key worker** |  |
| Yes | 11342 (22.1%) |
| **Chronic physical illness** |  |
| Yes | 19655 (38.2%) |
| **Mental health disorder** |  |
| Yes | 10219 (19.9%) |
| **Low social support** |  |
| Yes | 13235 (25.7%) |
| **Psychological/physical abuse** |  |
| Yes | 5798 (11.3%) |
| **COVID-19 symptoms** |  |
| Yes | 7618 (14.8%) |
| **Depressive symptoms (PHQ-9)** |  |
| Minimal/mild | 35715 (69.5%) |
| Moderate | 12451 (24.2%) |
| Severe | 3251 (6.3%) |
| **Note.** Weighted descriptive statistics of the sample at the first assessment. |  |
| <sup>a</sup> Quartiles were computed prior to weighting so actual percentages vary from 25% in the weighted sample. |  |

### Group-based depressive symptom trajectories

The LGM analysis resulted in three distinct DSTs (Figure 1): Class 1 – Low depressive symptoms [N=30,850 (60%)]; Class 2 – Moderate depressive symptoms [N=14,911 (29%)]; Class 3 – Severe depressive symptoms [N=5,656 (11%)], which decreased following the start of lockdown but began to increase again in weeks 5 and 6.

**Figure 1.**
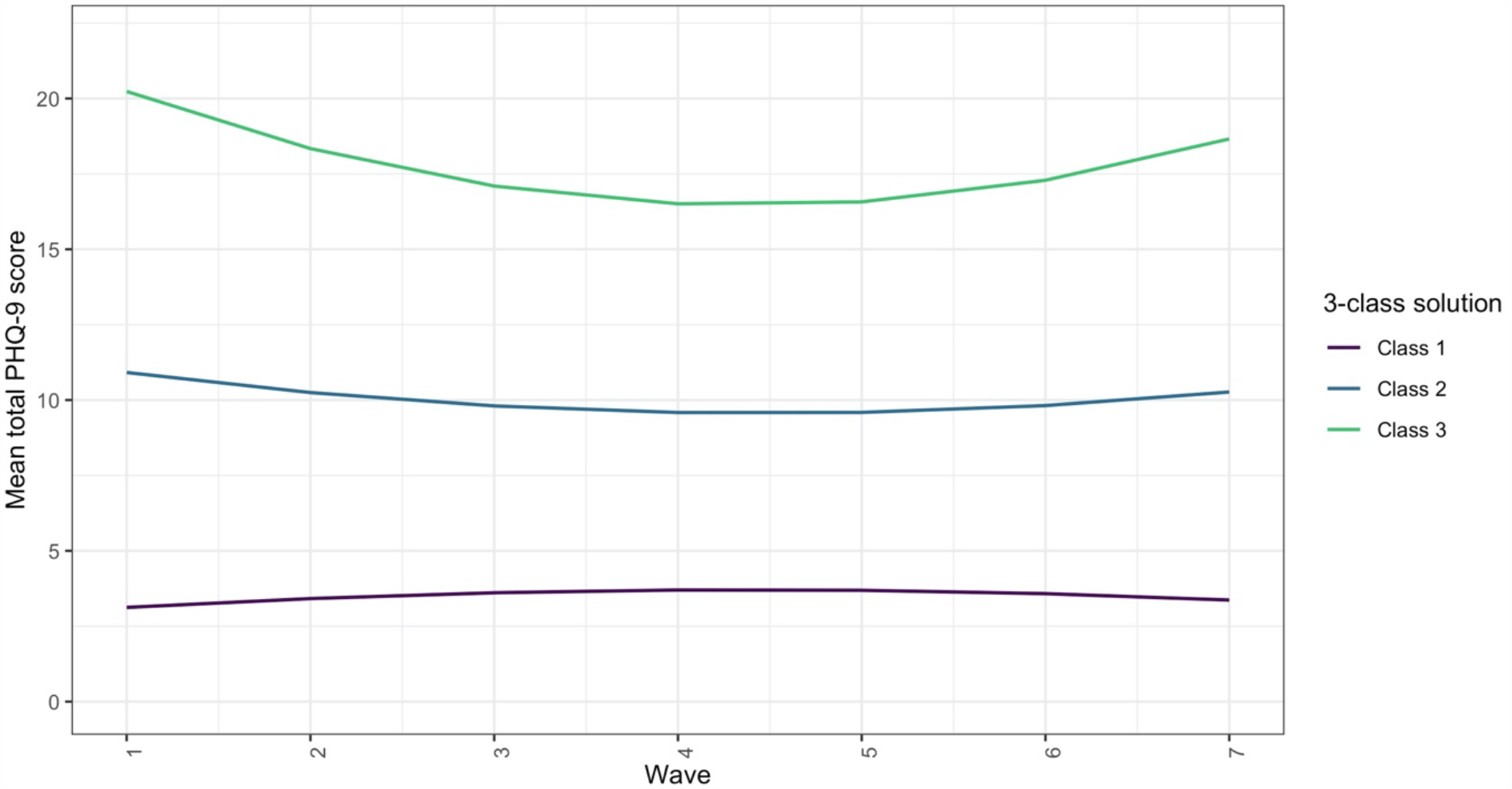
Group-based trajectories of depressive symptoms (wave 1-7) ***Note***. The Low Depressive symptom trajectory (Class 1) includes participants with minimal depressive symptoms at all waves [30,850 individuals (60%)]. The Moderate depressive symptom trajectory (Class 2) represents participants with moderate depressive symptoms throughout the study [14,911 individuals (29%)]; The Severe depressive symptom trajectory (Class 3) represents participants with persistently severe levels of depressive symptoms [5,656 individuals (11%)].

### Associations of sociodemographic, health-related, and psychosocial vulnerabilities with depressive symptom trajectories

The associations of the vulnerability factors with the DSTs are illustrated in Figure 2 and reported in sTable 5 (Supplement). In Model 1, adjusted for sex, age, and COVID-19 symptoms, low SEP was positively associated with both the moderate [OR=1.97(1.87;2.08)] and severe [OR=5.22(5.08;5.36)] DST. These results were slightly reduced but still maintained when adjusting for other vulnerability factors (Model 2). Non-white ethnicity was associated with a greater risk of moderate depressive symptoms [OR=1.21(1.03;1.40)] but not severe depressive symptoms [OR=1.07(0.85;1.28)]. However, these results were attenuated when adjusting for other vulnerabilities. Participants with key worker roles were less likely to experience severe depressive symptoms than those without such roles [OR=0.66(0.53;0.80)], but this result was not maintained when adjusting for other vulnerabilities.

**Figure 2.**
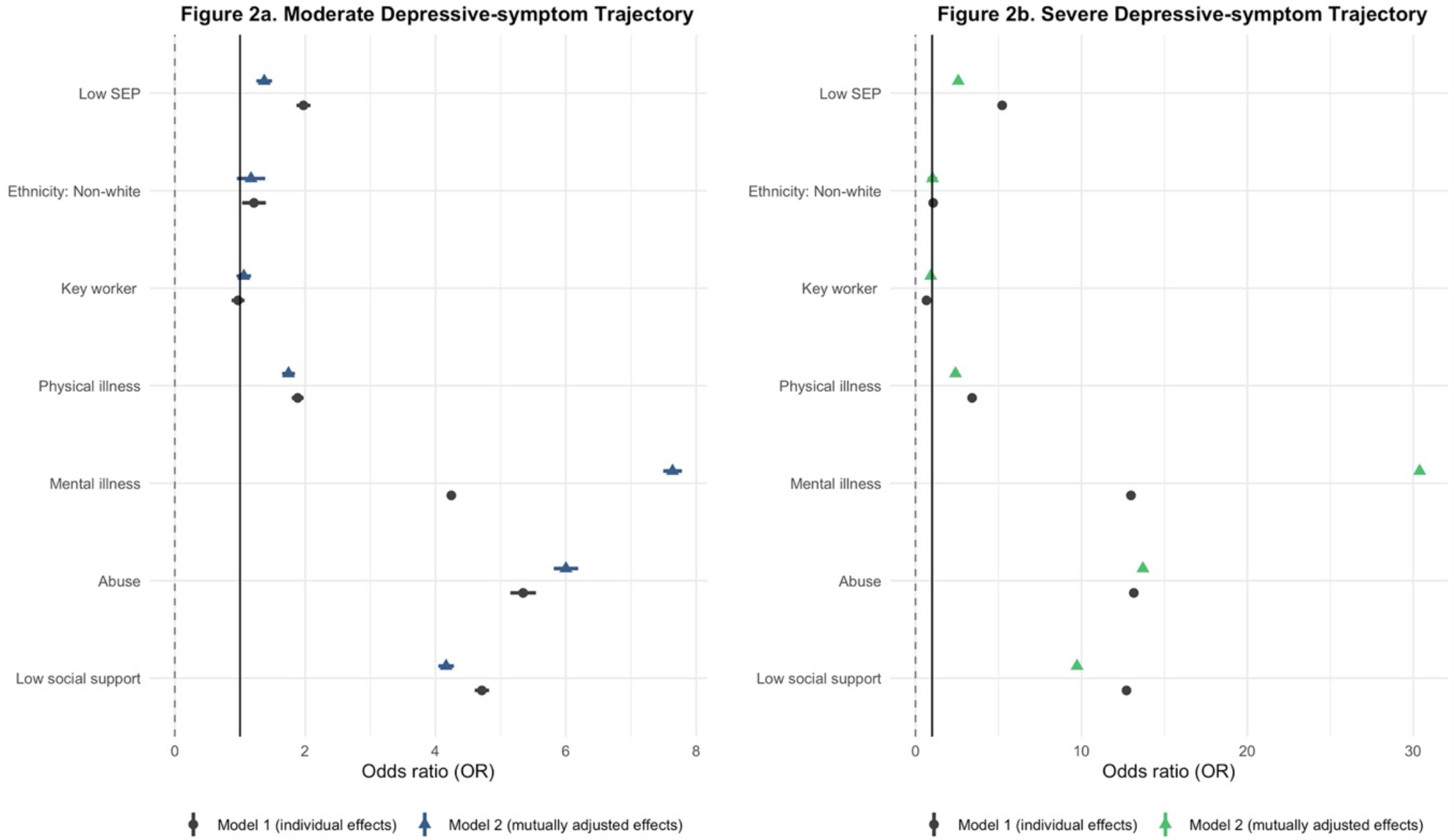
Associations of sociodemographic, psychosocial, and health-related vulnerabilities with the Moderate and Severe Depressive symptom trajectories. ***Note***. The odds ratios represent the risk of belonging to the Moderate or Severe Depressive symptom trajectory compared with the Low trajectory. All models were adjusted for sex, age, and COVID-19 diagnosis and weighted using survey weights.

Health-related vulnerabilities were related to a higher likelihood of moderate and severe DSTs. The largest ORs were observed for pre-existing mental health conditions [OR_Moderate_=4.24(4.24;4.24), OR_Severe_=13(12.87;13.10)], followed by physical health conditions [OR_Moderate_=1.89 (1.79;1.98), OR_Severe_=3.41(3.29;3.54)]. Psychosocial vulnerabilities were also linked to an elevated risk of moderate and severe depressive symptoms. The ORs of moderate depressive symptoms were 5.34(5.15;5.54) and 4.71(4.60;4.82) higher for participants who had experienced abuse and those with low social support respectively. In addition, the risk of severe depressive symptoms associated with abuse [OR=13.16(12.95;13.37)] and low social support [OR=12.72(12.57;12.86)] was more than twice as large as that of moderate depressive symptoms. These vulnerability factors all remained as risk factors when adjusting for other vulnerabilities.

## DISCUSSION

This is the first study to investigate trajectories of depressive symptoms among vulnerable groups in the UK during the COVID-19 pandemic using data from a large longitudinal study. Our analysis revealed a number of key findings. First, trajectories of depressive symptoms in the sample were relatively constant across the first 6 weeks of lockdown. However, this does not imply that mental health has not been affected by the pandemic. Data from various surveys suggest that mental health worsened in the lead up to lockdown being introduced in the UK, leading to higher than usual levels of anxiety and depression ^25–29^. Although our analysis does not claim to show prevalence due to the non-random nature of the sample, it is notable that the average scores presented here were substantially higher than PHQ-9 averages previously reported in population estimates in other high income countries such as the US ^30^. Further, our results suggest that there was little improvement in depression in the first few weeks of lockdown.

Our findings also showed that individuals facing certain sociodemographic, psychosocial and health-related vulnerabilities were at heightened risk of experiencing moderate and severe depression during lockdown. The risk of both moderate and severe depressive symptoms was considerably higher among people experiencing abuse, low social support, in individuals with low SEP, and in those with pre-existing mental and physical health conditions. Notably, these associations were larger than might be expected from previous literature, particularly for psychosocial vulnerabilities. For example, meta-analytical results suggest that the odds of depression are 1.87(1.42;2.46) higher in people exposed to intimate partner violence as compared to non-exposed people ^15^, and 0.74 (0.72;0.76) lower in people with high levels of social support compared with those with poor social support ^31^. The relationships of abuse and low social support with severe depressive symptoms were more than 5 times larger in our study, with odds ratios of 13.2 for abuse and 12.7 for low social support. Likewise, previous meta-analyses have shown that low SEP and chronic physical health conditions such as cardiovascular disease increase the odds of depression by 1.81(1.57;2.10) and 1.75(1.36;2.26) respectively. Our data suggest that the odds of severe depressive symptoms were 3.41 times higher for people with chronic physical illnesses and 5.22 times higher in those facing socioeconomic disadvantage. These figures are particularly worrying considering likely increases in rates of abuse, domestic violence, and unemployment and diminished access to social networks during the current COVID-19 pandemic ^3,32^. As such, the results presented here suggest that certain groups who usually experience higher odds of depression are at even greater risk during the current pandemic.

Notably, non-white ethnicity was related to higher depressive symptoms, but these results were explained by other sociodemographic characteristics, abuse and social support, as well as pre-existing physical or mental illnesses. Thus, it appears that the higher prevalence of other socio-economic and psychosocial vulnerabilities amongst ethnic minority groups is a greater risk factor than ethnicity itself for depression during the pandemic. Additionally, despite evidence suggesting that people with key worker roles such as healthcare workers are particularly susceptible to poor mental health outcomes during epidemics ^18^, our results show that the risk of elevated depressive symptoms was similar in participants with and without key worker roles. However, our key worker measure was not limited to healthcare professions but also included other keyworker roles such as teachers and transport workers who might be experiencing different levels of work-related stress during the current pandemic. Further, it is possible that self-selection bias determined these findings, with only key workers who are psychologically coping with the current demands of their roles taking the time to participate in the research. This study has a number of strengths, including the large sample size, the longitudinal study design, and the statistical methods employed to explore the link between specific vulnerabilities and DSTs. However, our findings need to be interpreted in light of several limitations. First, our sample, though well-stratified across socio-demographic characteristics and weighted to population proportions, was not random, and hence is not nationally representative. It is possible that the study inadvertently attracted individuals experiencing greater psychological distress during the pandemic, or individuals who are more engaged or interested in mental health. Hence, the results shown here are not presented as prevalence figures but are instead used to understand risk factors. Second, the data used in the present analyses are based on self-report measures, bearing the risk of self-report bias. Lastly, causality cannot be assumed since the study is observational and only provides information about the severity of depressive symptoms among vulnerable groups over time. As we lack data on individuals from prior to lockdown being brought in the UK, we are unable to identify if and how patterns of depressive symptoms during lockdown vary relative to individuals’ usual mental health.

In conclusion, our analysis suggests that certain vulnerable groups are at particular risk of experiencing elevated depressive symptoms during the current COVID-19 pandemic, including people with pre-existing mental and physical health conditions, experience of physical/psychological abuse, low social support, and those with low SEP. These groups may be experiencing even greater risk than in ordinary circumstances. In contrast, key workers and individuals from ethnic minority groups were not more likely to report depressive symptomatology when other vulnerabilities were taken into account. These differential associations highlight the importance of developing strategies to identify vulnerable individuals, reallocate mental health services to those in need, and provide evidence-based treatments to alleviate depressive symptoms.

## Data Availability

The anonymous data will be made publicly available after the end of the pandemic.

## DECLARATIONS

### Ethics approval and consent to participate

Ethical approval for the COVID-19 Social Study was granted by the UCL Ethics Committee. All participants provided fully informed consent. The study is GDPR compliant.

### Contributors

All authors contributed significantly to the conception, design, analysis or interpretation of data and were involved in revising it critically for intellectual context. The final submission of this paper was approved by all authors.

